# A Randomized Feasibility Trial of a Multicomponent Quality Improvement Strategy for Chronic Care of Cardiovascular Diseases: Findings from the C-QIP Trial in India

**DOI:** 10.64898/2026.01.28.26345028

**Authors:** Kavita Singh, Ambuj Roy, Dimple Kondal, Kalyani Nikhare, Mareesha Gandral, Satish G Patil, Kiran Aithal, MP Girish, Mohit Gupta, Kushal Madan, JPS Sawhney, Kamar Ali, Meetushi Jain, Savitesh Kushwaha, Devraj Jindal, Emily Mendenhall, Shivani A. Patel, KM Venkat Narayan, Nikhil Tandon, Mark D Huffman, Dorairaj Prabhakaran

**Affiliations:** Public Health Foundation of India, New Delhi, India; Heidelberg Institute of Global Health, Heidelberg University, Germany; Centre for Chronic Disease Control, New Delhi, India; All India Institute of Medical Sciences, New Delhi, India; SDM College of Medical Sciences and Hospital, Karnataka, India; GB Pant Hospital, New Delhi, India; Sir Ganga Ram Hospital, New Delhi, India; Georgetown University, Washington, D.C., USA; Emory Global Diabetes Research Center of Woodruff Health Sciences Center and Emory University, Atlanta, USA; Washington University School of Medicine, St. Louis, USA; The George Institute for Global Health, University of New South Wales, Sydney, Australia

**Author notes:** **Corresponding author** Kavita Singh Senior Research Scientist Public Health Foundation of India, and Centre for Chronic Disease Control, New Delhi, India Heidelberg Institute of Global Health, Heidelberg University Hospital, Im Neuenheimer Feld 130.3, 69120 Heidelberg. senior co-author (Mark D Huffman, Dorairaj Prabhakaran).

## Abstract

**Background:** Chronic cardiovascular diseases (CVD) care quality remains suboptimal, globally. This study evaluated the feasibility and preliminary effect of a multicomponent, collaborative quality improvement (C-QIP) strategy among patients with CVD attending outpatient clinics in India.

**Methods and Findings:** We conducted a pragmatic feasibility randomized controlled trial in patients with ischemic heart disease, ischemic stroke or heart failure across public and private hospitals in India. Participants were individually randomized to C-QIP strategy (electronic decision support system, eDSS for providers, task-sharing with non-physician health workers, patient education, and SMS text reminders, and audit-feedback) or usual care. The primary outcomes were implementation measures: feasibility, fidelity, adoption, and acceptability from provider’s and patient’s perspectives. Secondary outcomes included prescription of guideline-directed medical therapy (GDMT), adherence to prescribed therapy, processes of care, and CVD risk factors. Of 410 participants enrolled (intervention arm=206 and usual care arm=204), mean age was 57.5 years, and 73.0% were male. Prior history of coronary heart disease was 74.6%, ischemic stroke: 18.5%, and heart failure: 18.0%. At trial end (mean follow-up 18 months), implementation outcomes were strong: retention at end-of-study was 192/206 (93.2%) in C-QIP and 187/204 (91.7%) in usual care arm; fidelity of the intervention remained high, e.g., 187/198 (94.4%) patients received lifestyle advice at end-of-study. Clinician adoption of eDSS prompts was high, and acceptance of DSS prompts varied by type of prompts, and both patients and providers reported high acceptability at trial end. GDMT use improved significantly in C-QIP vs usual care arm at end-of-study: in patients with ischemic heart disease use of antiplatelet + statin + ACEi/ARB + beta-blocker was 58.3% vs 32.4%, RR=1.45 (95%CI: 1.18–1.78); and among patients with ischemic stroke use of antiplatelet + statin + ACEi/ARB or diuretic was 76.7% vs 31.8%, RR=2.41 (95%CI: 1.52–3.81). GDMT among patients with heart failure were not different between groups (e.g., ACEi/ARB /ARNI + beta-blocker + MRA, 48.9% vs 48.6%, RR=1.26, 95%CI: 0.82–1.94). Patient adherence to prescribed therapy improved in C-QIP vs usual care arm: medications 90.9% vs 82.3%, RR 1.08 (1.04–1.12); diet plan 91.9% vs 82.3%, RR 1.07 (1.02–1.13); and physical activity 91.4% vs 70.4%, RR=1.23 (95%CI: 1.16–1.30). Processes of care improved significantly in C-QIP vs usual care arm, including more structured reminders (e.g., call after missed appointment 70.7% vs 4.4%, p<0.001) and longer clinician contact time (median 10 vs 7 minutes, p<0.001). CVD risk factors showed small, non-significant trends (e.g., modest diastolic BP reduction) for between-group differences in blood pressure, lipids and glycemia.

**Conclusions:** The C-QIP trial demonstrated that a multicomponent strategy is feasible, acceptable, and improved processes of chronic CVD care in India. Future large, confirmatory hybrid trials are needed to establish whether such quality improvement strategies can reduce cardiovascular morbidity and mortality.

**Trial Registration:** Clinicaltrials.gov number: NCT05196659

Clinical Trials Registry India: CTRI/2022/04/041847

## Background

Cardiovascular diseases (CVD) caused an estimated 20.5 million deaths and 300 million disability-adjusted life years in 2024[1, 2]. The societal and economic consequences are profound, particularly in low-and middle-income countries (LMICs), where CVD affects working-age adults[3, 4]. Secondary prevention is a critical strategy to reduce the rapidly growing CVD burden[5]. Despite advances in evidence-based guidelines, wide gaps persist in their implementation and quality of chronic CVD care across diverse healthcare settings[6, 7]. For instance, the Prospective Urban Rural Epidemiology (PURE) study underscores the magnitude and persistence of this gap among adults with prior CVD across 17 countries, use of proven secondary prevention drugs was strikingly low overall: antiplatelets 25%, β-blockers 17%, ACEi/ARB 20%, statins 15%, with the lowest use in low-income settings (e.g., statins 3%)[8]. Updated 2025 PURE analyses reported global disparities in CVD drug use over a median follow-up of 12 years and found persistently low and declining use of secondary prevention drugs among people with CVD[9]. Globally, use of ≥1 recommended class (antiplatelet, statin, ACEi/ARB, β-blocker) was ∼41% at baseline in 2007, peaked near 43%, then fell to ∼31% by the last visit (2019); in high-income countries it decreased from ∼89% to ∼77%. In India, use of ≥1 secondary-prevention medication class was 21.4% at baseline, rose to a peak of 43.0% by visit 4 (mid-period), and then declined to 21.6% by the last visit; the highest use of ≥3 classes reached 10.7% during follow-up. These data on low use of CVD secondary prevention drugs underscore a stalled (or worsening) implementation gap, especially in lower-resource settings [9].

These gaps further reflect multilevel barriers, including fragmented healthcare delivery, clinical inertia, inadequate universal healthcare coverage, limited patient awareness, and poor adherence to long-term treatment[10, 11]. Closing these persistent “know-do” gaps requires pragmatic, context-specific strategies that go beyond single interventions[12, 13]. To address these gaps, the Indian government has updated National Programme for Prevention and Control of Noncommunicable Diseases (NP-NCD, 2023 operational guidelines)[14], with emphasis on integrated team-based care, use of digital health tools, task-sharing with non-physician health workers, and strengthening secondary prevention services across public and private systems[15]. In light of this evolving policy context, the C-QIP trial builds on prior implementation trials in India, including, the ACS-QUIK trial[16, 17], focused on hospital-based quality improvement intervention in patients with acute coronary syndrome, the SPREAD[18] trial involving community health worker led intervention to support patients after acute coronary syndrome, and the CARRS Translation Trial[19, 20] utilizing multicomponent, technology enabled-intervention for cardiovascular risk reduction in people with poorly controlled type 2 diabetes.

However, strategies that combine system-, provider-, and patient-level interventions that integrates clinical decision support, task-sharing, patient engagement, and audit-feedback to improve the quality of chronic CVD care have not been systematically tested in Indian context. To address this critical gap, we evaluated the feasibility and preliminary effect of a collaborative quality improvement (C-QIP) strategy compared with usual care among patients with CVD attending outpatient clinics in India.

## Methods Study design

The C-QIP trial was a multi-site, individual-level randomized (1:1), controlled, open label, feasibility trial with collection of implementation and effectiveness outcomes to inform a larger outcomes-driven hybrid effectiveness-implementation trial. Full details of the study design and methods have been described previously[21] (and trial protocol provided in **Supplement 1**). The trial protocol was registered prospectively on Clinicaltrials.gov: <u>NCT05196659</u> and Clinical Trials Registry India: <u>CTRI/2022/04/041847</u>. The C-QIP trial was conducted at four hospitals (2 public and 2 private) located in Delhi and Karnataka in India. The C-QIP trial began recruitment from 12 September 2022 and completed target recruitment in 30 September 2023. Follow-up visits were completed in November 2024.

### Participants

Patients with a confirmed diagnosis of CVD including ischemic heart disease, ischemic stroke, or heart failure irrespective of ejection fraction were eligible for the trial. Patients aged ≥18 years, both sexes, living within same city/town as one of the sites and those willing to provide consent were recruited in the trial. Pregnant women and patients having disease associated with frequent hospitalization (e.g., advanced cancer, end stage renal disease); bedridden or debilitating conditions, and life expectancy less than 12 months were excluded from the trial. The recruitment goal was 100 participants at each of the four clinic sites for a total sample size of 400 participants to ensure feasibility, and balanced representation across diverse outpatient settings in India. The site-level target of 100 participants facilitated operational efficiency, consistent delivery of the intervention, and evaluation of between-site heterogeneity in implementation and clinical outcomes. Eligible participants attending the outpatient clinic at the four sites were recruited prospectively. Participants provided written informed consent and were then randomly allocated to one of the study arms.

### Randomization

Randomization occurred via a computerized randomization program that was accessed through a secure web interface. The random allocation sequence was in a uniform 1:1 allocation ratio with a varied block size of 4, 6 and 8 and concealed from study personnel. Study staff enrolled/randomized eligible patients by entering data into the secure web interface.

### Intervention

A multicomponent C-QIP strategy was developed using a co-design approach engaging with multiple stakeholders (patients living with heart disease, and their caregivers, non-physician health workers [e.g., nurses, pharmacists], physicians, cardiologists, healthcare administrators, and policy makers). The co-design process involved i) a systematic scoping review of published literature[22], ii) qualitative interviews with patients, caregivers, providers, healthcare administrators, and policy makers[23], and iii) a modified Delphi survey among experts practicing cardiology or related fields to generate consensus around the C-QIP strategy components[24]. The C-QIP strategy consisted of *five components* - 1) Trained and supervised non-physician health worker (named as cardiovascular care coordinator) facilitated care for patients with CVD.

The cardiovascular care coordinator managed around 50-60 patients randomized to the intervention arm and assisted the physician to devise a patient-tailored CVD care plan based on patient’s CVD risk factor levels, adherence, and motivation. 2) Electronic health record – decision support system (EHR-DSS)[21] to improve physician’s responsiveness to timely treatment modification for CVD management. The DSS provided automated decision-support prompts of guideline-recommended care tailored to each patient’s CVD condition, risk level, and adherence to current medications. The EHR-DSS was co-managed by the site physician and cardiovascular care coordinator. The cardiovascular care coordinator collected and entered all patient related data into the EHR. The site physician reviewed the computer generated DSS prompts (CVD management plan) and made appropriate modifications to the treatment plan. 3) Text-message based reminders tailored to patient’s lifestyle habits and to promote healthy behaviors and regular clinic visit attendance and laboratory testing, and for improving adherence to prescribed therapy. 4) Patient diary containing reinforcement tool for lifestyle modifications and visual tool for medication adherence (VITA), 5) Quarterly audit and feedback reports for providers. The audit and feedback reports were delivered by email to the participating site clinical team. Audit/feedback report included patient-level profiles and aggregate summaries of key quality indicators (use of aspirin, ≥2 blood pressure–lowering medicines, statins, and glucose-lowering medicines for patients with diabetes), as well as clinic-level averages of systolic/diastolic blood pressure, LDL cholesterol, and glycated hemoglobin (HbA1c). Site teams were instructed to use this information within a Plan–Do–Study–Act (PDSA) cycle, reviewing performance against quarterly goals, identifying gaps, and implementing corrective actions. (see online **supplement 2** for details on C-QIP strategy components).

### Control arm: Usual Care

Participants randomized to the control arm received a leaflet (i.e., printed information) on healthy behaviors to prevent recurrent CVD events, delivered by a different site team member to minimize the risk of contamination. Patients in the usual care group received routine care and were only contacted once per year before their scheduled annual study visit.

### Trial procedures

The first study visit included screening, randomization, and baseline assessment including clinical measures (i.e., blood pressure, heart rate, weight, body mass index). Further, fasting blood glucose (FBG), HbA1c, and lipids were tested for all randomized participants at baseline. In addition, validated questionnaires were used for the assessment of processes of care measures including self-care, adherence to medications, diet plan and physical activity, global health assessment scale for physical and mental health assessment, patient’s treatment satisfaction, health service use, and reminders for clinic visit or lab tests [21].

Participants randomized to the intervention arm had more frequent follow-up visits per the DSS recommendations, for e.g., in case of uncontrolled blood pressure (BP: ≥140/90 mm Hg), or high-risk heart failure patients. Intermediate visit assessment for intervention arm participants included interval history, presenting complaints, self-care measures, and vital signs. All intermediate visits data were documented in the EHR. The post-randomization follow-up period ranged from 12 - 24 months, depending on participant enrolment in the trial. At annual visit and end of study visit, all participants were assessed for study outcomes, including implementation outcomes, processes of care measures and clinical measures including BP, lab tests, self-care behaviors, health-related quality of life, treatment satisfaction, consultation time, reminders system for next clinic visit/lab tests, and health service utilization including outpatient and inpatient care.

### Outcomes

The primary implementation outcomes were feasibility, fidelity, adoption and acceptability of the multicomponent C-QIP strategy from providers and patient’s perspectives, and details of data sources, timing and success thresholds were prespecified in the published trial protocol [21]. *Feasibility* was defined as the extent to which C-QIP strategy was successfully implemented in routine outpatient workflows with available staff, time and infrastructure (e.g., enrollment of eligible patients and retention ≥80%). *Fidelity* was defined as the degree to which C-QIP components were delivered per protocol (e.g., cardiovascular care coordinator delivered counseling for adherence to GDMT, lifestyle advice, patient diary and VITA tools: ≥80%; SMS sent/delivered: ≥80%; A&F report delivered/reviewed ≤7 days with ≥1 change tested per cycle). *Adoption* was defined as the initial uptake of C-QIP DSS prompts by clinicians (DSS used in eligible visits: ≥70%; clinicians using DSS: ≥80% of enrolled patients; clinics assigning NPHW roles (100%); patients consenting to SMS/VITA tools (≥80%)). *Acceptability* was defined as perceived satisfaction, fit, and value among providers and patients (≥80% of providers and patients willing to continue using C-QIP strategy). Secondary outcomes included processes of care measures (e.g., prescription of GDMT, and patient’s adherence to prescribed medications, diet plan, physical activity, clinic visits attendance, routine monitoring of CVD risk factors, mean changes in BP, LDLc, and HbA1c, reminders for clinic visit/lab test, consultation time, and family support for self-care) from baseline to end of study visit. GDMT for ischemic heart disease was defined as antiplatelet + statin + Beta blocker + Angiotensin-Converting Enzyme inhibitor / Angiotensin II Receptor Blocker (ACEi/ARB). GDMT for ischemic stroke was defined as antiplatelet + statin + ACEi or ARB / Diuretic. GDMT for heart failure was defined as ACEi or ARB /ARNI + Beta blocker + Mineralocorticoid Receptor Antagonist (MRA) + Sodium–Glucose Co-Transporter 2 inhibitor (SGLT2i).

### Statistical analysis

The sample size proposed for this feasibility trial was 400 patients randomized from four sites (100 patients per site). Given the primary goal of the trial was to establish that data collection protocols were feasible, fidelity of implementation, and participant adherence and retention, and to estimate the confidence intervals (CIs) for processes of care measures and clinical outcomes, a sample size of at least 70 per arm was considered adequate[25, 26]. We proposed to recruit 100 participants per site (taking into account 10% drop-out rate) with a total of 400 participants to demonstrate the feasibility of C-QIP strategy implementation. The baseline characteristics of the participants were reported by treatment group. The results for the primary implementation outcomes i.e., feasibility, fidelity, adoption, and acceptability from provider’s and patient’s perspectives were reported as number (percentages) over the study period in the intervention arm only. For instance, the number (percentage) for the acceptance of clinical DSS prompts by the treating doctors was reported overall and stratified by sites.

The analyses for secondary outcomes such as prescription of medications (provider-level), adherence to prescribed treatment, mean changes in BP, LDLc, and HbA1c (patient-level outcomes) were conducted with the intention to treat (ITT) principle based on the pre-specified statistical analysis plan (**Supplement 3**). The analysis for the prescription of medications, and adherence to self-care practices, was conducted using generalized estimating equations (GEE) using log binomial model taking into account correlation of observations within participants over time. The covariates included in the model were time (baseline and EOS), and treatment group (C-QIP strategy and usual care). The model was adjusted for the age, sex, and site. We used the Poisson link if the model did not converge using binomial link. Effect sizes (relative risk, RR or risk difference) with 95% CIs were reported.

The analysis for the continuous outcomes: processes of care, health status, treatment satisfaction, clinical measure (SBP, DBP, LDLc, HbA1c, serum creatinine, weight and body mass index, BMI) was conducted using linear regression with GEE model. The covariates included in the model were time (baseline and EOS), treatment arm (C-QIP strategy and usual care), and adjusted for age, sex and site. The beta coefficient with 95% CI was reported for continuous outcomes. We performed the ITT analysis and did not impute the missing continuous outcomes. We compared the baseline characteristics between the final study sample and those who were lost to follow-up.

Subgroup analyses were conducted for self-reported medication adherence and systolic blood pressure as these measures were applicable to all patients enrolled in the study (irrespective of type of CVD condition). The prespecified subgroups were age; sex; facility type; presenting condition (ischemia heart disease, ischemic stroke, heart failure); prior history of hypertension, diabetes; baseline BMI, baseline BP; and LDLc. For each of the aforementioned subgroups, we repeated the GEE analysis with the addition of the subgroup variable along with its interaction with treatment group. Heterogeneity was assessed based on the significance of the interaction term. Serious adverse events including deaths, and hospitalizations were reported overall and by treatment group. Data analysis was performed using STATA 18.0 and a p-value of <0.05 was considered for statistical significance.

### Ethics

The C-QIP Trial protocol was reviewed and approved by the institutional ethics committee of the Public Health Foundation of India (TRC-IEC-382.2/18) and Health Ministry Screening Committee in India (2018-0491). All participating sites also obtained their respective local site ethics committee approvals. Participation in this trial was voluntary, and a signed consent form was obtained from each trial participant.

## Results

### Feasibility and participant flow

A total of 492 participants were screened, of whom 410 were randomized to either the C-QIP strategy (n=206) or usual care (n=204). Baseline assessments were completed for 198=C-QIP and 203=usual care participants, with BP, LDL-C, and fasting glucose/HbA1c measurements available in over 90% of participants. At 12 months, 91/114 expected patient follow-up visits were completed in the C-QIP arm and 97/129 in usual care, corresponding to 85% visit completion. By the end of study (EOS), 192 participants in the C-QIP and 187 in usual care arm completed final study visit, yielding retention rates of 93.2% and 91.7%, respectively. The median follow-up duration was similar between arms, i.e., 15.3 months (interquartile range, IQR 12.8–19.8) in C-QIP and 15.1 months (IQR 12.6–20.1) in usual care arm, equivalent to roughly 459 vs 452 days. Dropouts and deaths were few (11 vs 1 dropouts; 9 vs 10 deaths in the C-QIP and usual care arm, respectively), confirming high trial feasibility and participant retention across both public and private sites (**Figure 1**).

**Figure 1:**
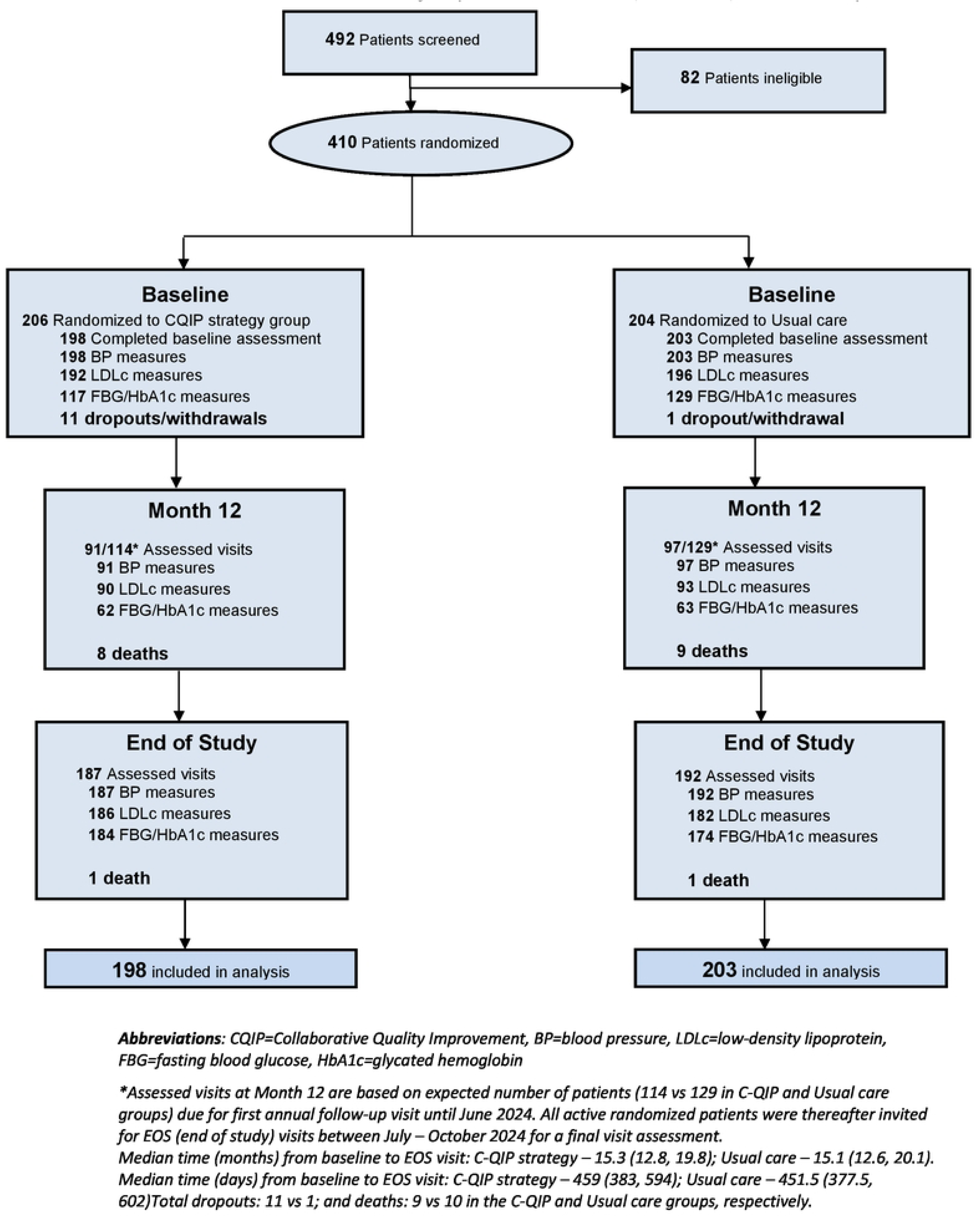
Trial Profile: Feasibility of patient recruitment, retention, and follow-ups

### Baseline characteristics by treatment group

The C-QIP strategy (n=198) and usual care arm (n=203) were well-matched at baseline. The mean age was 57.4 years (SD 11.7), with similar proportions of male participants (72.7% in C-QIP vs. 73.9% in usual care) and mean (SD) years of education: 10.5 (5.7) years in C-QIP vs 11.0 (5.3) in usual care arm. Prior history of CVD including coronary heart disease (76.3% vs 71.4%), heart failure (22.7% vs 18.2%), and ischemic stroke (15.2% vs 21.7%), were not significantly different between C-QIP and usual care arms, respectively. Baseline CVD risk factors were also not different between the two groups: mean SBP 126.4 ± 19.8 mmHg vs 129.2 ± 16.5 mmHg; DBP 75.4 ± 12.1 vs 77.2 ± 10.9 mmHg; LDL-C 82.1 ± 40.1 mg/dL vs 78.8 ± 34.3 mg/dL in the C-QIP vs usual care arm, respectively (**Table 1**).

**Table 1:** Baseline characteristics of study participants.

|  | C-QIP strategy | Usual care |
| --- | --- | --- |
|  | N=198 | N=203 |
| Age (years; mean, SD) | 57.2 (11.2) | 57.6 (12.2) |
| Male, n (%) | 144 (72.7%) | 150 (73.9%) |
| Education, (years; mean, SD) | 10.5 (5.7) | 11.0 (5.3) |
| Household Income (monthly) Indian rupees, median (IQR) | 20000<br>(10000, 30000) | 20000<br>(12000, 30000) |
| **Medical history** |  |  |
| Coronary heart disease, n (%) | 151 (76.3%) | 145 (71.4%) |
| Heart failure, n (%) | 45 (22.7%) | 37 (18.2%) |
| Ischemic stroke, n (%) | 30 (15.2%) | 44 (21.7%) |
| Hypertension, n (%) | 86 (43.4%) | 107 (52.7%) |
| Diabetes, n (%) | 82 (41.4%) | 86 (42.4%) |
| **Biomarkers** |  |  |
| Systolic BP (mm Hg; mean, SD) | 126.4 (19.8) | 129.2 (16.5) |
| Diastolic BP (mm Hg; mean, SD) | 75.4 (12.1) | 77.2 (10.9) |
| Total cholesterol (mg/dL; mean, SD) | 144.0 (46.5) | 145.2 (41.2) |
| LDLc (mg/dL; mean, SD) | 82.1 (40.1) | 78.8 (34.3) |
| FBG (mg/dL; mean, SD) | 123.2 (54.7) | 121.3 (55.3) |
| HbA1c (%; mean, SD) | 6.8 (1.5) | 6.7 (1.7) |
| **Medications use** |  |  |
| Antiplatelet | 178 (89.9%) | 173 (85.2%) |
| Lipid lowering (Statin) | 181 (91.4%) | 163 (80.3%) |
| ACEi/ARB | 123 (62.1%) | 116 (57.1%) |
| ARNI (HF=82) * | 14 (31%) | 11 (30%) |
| Beta-blockers | 148 (74.7%) | 136 (67.0%) |
| Calcium Channel Blockers | 36 (18.2%) | 35 (17.2%) |
| Diuretics | 16 (8.1%) | 14 (6.9%) |

**Abbreviations:** LDLc = Low-density lipoprotein cholesterol; FBG = Fasting Blood Glucose; HbA1c = glycated hemoglobin; ACEi = Angiotensin-Converting Enzyme Inhibitors; ARB = Angiotensin Receptor Blocker; ARNI = Angiotensin Receptor-Neprilysin Inhibitor; CCBs = Calcium Channel Blocker Values reported are mean (SD) or median (p25, p75) or n (%); p25 and p75 is the 25th and 75th percentile; \*#only reported for heart failure patients.

### Implementation outcomes: Fidelity, Adoption, and Acceptability

Implementation fidelity was high. Cardiovascular care coordinators (CCCs) delivered lifestyle counselling advice to 99.5% of participants at baseline, 79.8% at 12 months, and 94.4% at EOS (**Figure 2a**). At the final visit, counselling was provided to 99.5% participants for healthy diet, 98.4% for exercise, 90.9% for stress avoidance, and 90.4% for home BP monitoring. Among those eligible, counselling for smoking cessation reached 60% and alcohol moderation 33% at EOS (**Table S.1.**). Adoption of the eDSS was universal, i.e., 100% of follow-up visits included review of DSS prompts with variable acceptance across prompt types (antiplatelet, BP, lipid, glycemia) as illustrated in **Figure 2b and Figure S.1**. Intermediate-visit completion as per DSS recommendations between baseline and annual follow-up exceeded 90% for the first three quarterly visits and gradually declined thereafter (**Table S.2.**). Both patients and providers reported high acceptability at EOS as most participants expressed satisfaction with the C-QIP strategy and willingness to continue beyond the trial duration, while providers endorsed the DSS as helpful for guideline-based prescribing (**Figure 2c**). Among patients, 98.4% reported C-QIP made medication adherence easier, and 97.9% wanted to continue using C-QIP beyond trial duration. Among providers, 98.4% found DSS-supported care satisfactory, and 100% found it easy to follow guidelines (**Figure 2c**: Acceptability of C-QIP strategy).

**Figure 2:**
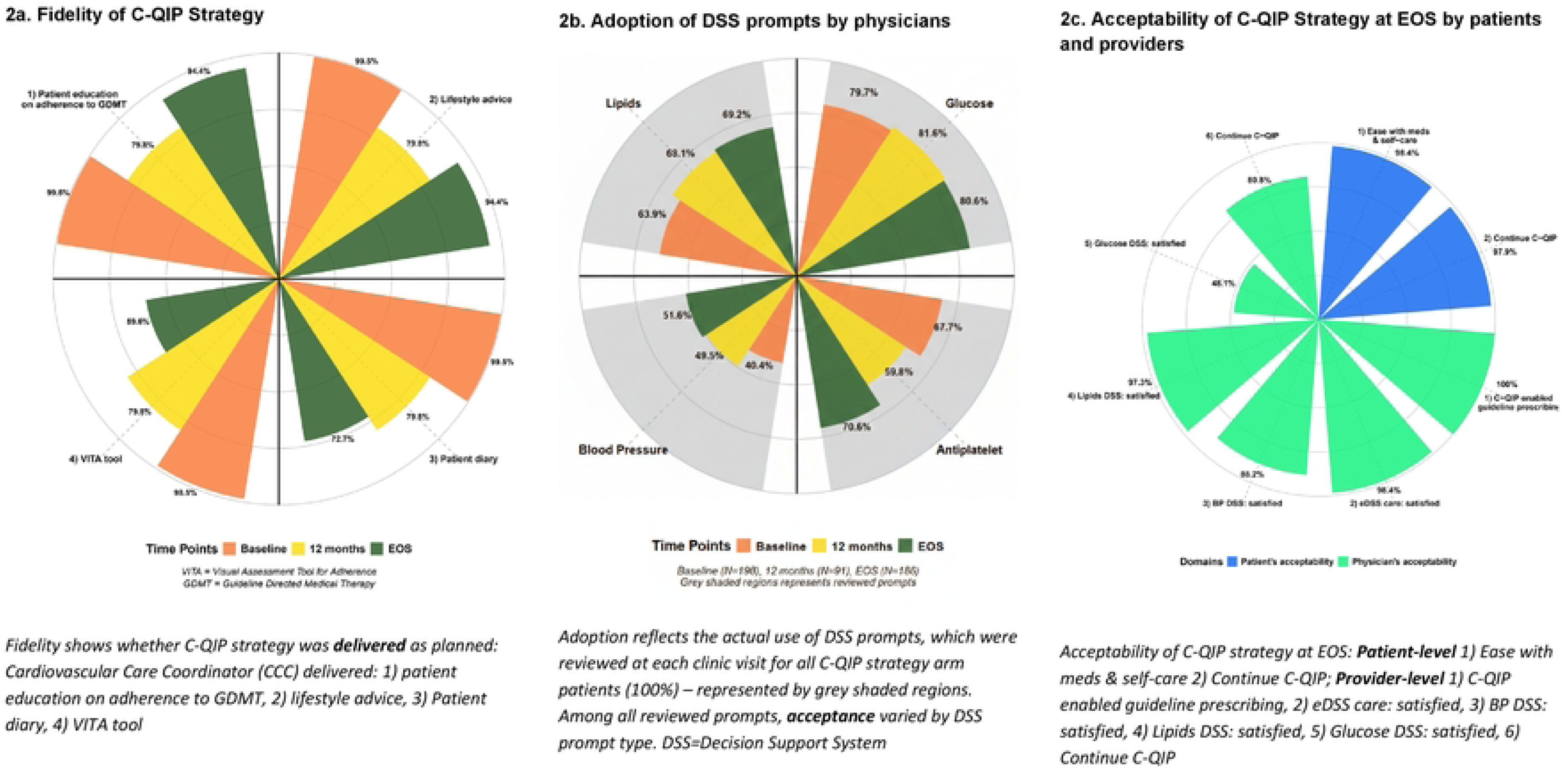
Implementation outcomes: Fidelity, Adoption, and Acceptability of C-QIP strategy

### Processes of care outcomes: prescription of GDMT, and adherence to therapy

At EOS, C-QIP increased prescription of antiplatelets, statins, ACEi/ARB, β-blockers, and diuretics, without differences for CCB or ARNI; evidence-based drug bundles improved significantly for patients with coronary heart disease (58.3% vs 32.4%; RR 1.45, 1.18–1.78) and ischemic stroke (76.7% vs 31.8%; RR 2.41, 1.52–3.81), but not for patients with heart failure (**Table 2**). Patient behaviors also favored C-QIP: adherence to medications (90.9% vs 82.3%; RR 1.08, 1.04–1.12), diet (91.9% vs 82.3%; RR 1.07, 1.02–1.13), and physical activity (91.4% vs 70.4%; RR 1.23, 1.16–1.30) was higher in the C-QIP vs usual care arm.

**Table 2:** Prescription of medications and adherence to self-care measures at end of study visit.

| Medications use | N | C-QIP strategy | N | Usual care | Relative risk (95% CI)* | p value | Risk difference % (95% CI)* |
| --- | --- | --- | --- | --- | --- | --- | --- |
| **Among all patients** |  |  |  |  |  |  |  |
| Antiplatelet | 198 | 176 (88.9%) | 203 | 154 (75.9%) | 1.11 (1.04, 1.19) | 0.003 | 9.0 (3.2, 14.8) |
| Lipid lowering | 198 | 178 (89.9%) | 203 | 148 (72.9%) | 1.18 (1.10, 1.27) | <0.001 | 14.2 (8.2, 20.2) |
| ACEi/ARB | 198 | 127 (64.1%) | 203 | 100 (49.3%) | 1.17 (1.01, 1.34) | 0.032 | 9.2 (0.9, 17.5) |
| Beta-blockers | 198 | 148 (74.7%) | 203 | 124 (61.1%) | 1.17 (1.05, 1.29) | 0.004 | 11.0 (3.7, 18.3) |
| Calcium channel blockers | 198 | 42 (21.2%) | 203 | 37 (18.2%) | 1.12 (0.78, 1.60) | 0.545 | 2.2 (-4.8, 9.1) |
| Diuretics | 198 | 26 (13.1%) | 203 | 11 (5.4%) | 1.82 (1.02, 3.28) | 0.045 | 5.0 (0.2, 9.8) |
| ARNI# | 45 | 14 (31.1%) | 37 | 11 (29.7%) | 1.03 (0.60, 1.77) | 0.919 | 0.9 (-16.3, 18.1) |
| **Patients with coronary heart disease (N=296)** |  |  |  |  |  |  |  |
| Medications use | N | C-QIP strategy | N | Usual care | Relative risk (95% CI)* | p value | Risk difference % (95% CI)* |
| Antiplatelet | 151 | 141 (93.4%) | 145 | 119 (82.1%) | 1.08 (1.02, 1.14) | 0.007 | 7.0 (2.0, 12.) |
| Lipid lowering | 151 | 143 (94.7%) | 145 | 117 (80.7%) | 1.14 (1.07, 1.21) | <0.001 | 11.8 (6.4, 17.1) |
| ACEi/ARB | 151 | 98 (64.9%) | 145 | 74 (51%) | 1.10 (0.94, 1.29) | 0.220 | 5.9 (-3.4, 15.3) |
| Beta-blockers | 151 | 128 (84.8%) | 145 | 99 (68.3%) | 1.17 (1.06, 1.30) | 0.003 | 12.5 (4.6, 20.4) |
| Antiplatelet + Lipid lowering drug (statin) + ACEi/ARB + Beta Blocker | 151 | 88 (58.3%) | 145 | 47 (32.4%) | 1.45 (1.18, 1.78) | <0.001 | 17.0 (8.1, 25.9) |
| **Patients with ischemic stroke (N=74)** |  |  |  |  |  |  |  |
| Medications use | N | C-QIP strategy | N | Usual care | Relative risk (95% CI)* | p value | Risk difference % (95% CI)* |
| Antiplatelet | 30 | 27 (90%) | 44 | 32 (72.7%) | 1.14 (0.96, 1.36) | 0.146 | 10.8 (-3.3, 25.0) |
| Lipid lowering | 30 | 27 (90%) | 44 | 28 (63.6%) | 1.34 (1.08, 1.67) | 0.009 | 23.4 (7.4, 39.4) |
| ACEi /ARB/Diuretics | 30 | 23 (76.7%) | 44 | 21 (47.7%) | 1.52 (1.11, 2.08) | 0.009 | 28.0 (7.8, 48.3) |
| Antiplatelet + Lipid lowering drug (statin) + ACEi/ARB/Diuretic | 30 | 23 (76.7%) | 44 | 14 (31.8%) | 2.41 (1.52, 3.81) | <0.001 | 44.3 (24.9, 63.7) |
| **Patients with Heart failure (N=82)** |  |  |  |  |  |  |  |
| Medications use | N | C-QIP strategy | N | Usual care | Relative risk (95% CI)* | p value | Risk difference % (95% CI)* |
| ACEi/ARB/ARNI | 45 | 33 (73.3%) | 37 | 25 (67.6%) | 1.16 (0.92, 1.47) | 0.209 | 11.0 (-6.0, 28.0) |
| Beta-blockers | 45 | 35 (77.8%) | 37 | 29 (78.4%) | 1.06 (0.87, 1.28) | 0.574 | 4.5 (-11.2, 20.2) |
| MRA | 45 | 26 (57.8%) | 37 | 25 (67.6%) | 1.16 (0.92, 1.46) | 0.198 | 9.9 (-4.9, 24.6) |
| SGLT2i | 45 | 19 (42.2%) | 37 | 16 (43.2%) | 1.28 (0.85, 1.91) | 0.237 | 11.6 (-7.2, 30.4) |
| ACEi /ARB/ ARNI + Beta blocker + MRA | 45 | 22 (48.9%) | 37 | 18 (48.6%) | 1.26 (0.82, 1.94) | 0.299 | 9.7 (-8.3, 27.7) |
| ACEi/ARB/ARNI + Beta blocker + MRA + SGLT2i | 45 | 12 (26.7%) | 37 | 11 (29.7%) | 1.27 (0.72, 2.25) | 0.411 | 6.9 (-9.5, 23.3) |
| **PATIENT-LEVEL MEASURES (self-reported)** |  |  |  |  |  |  |  |
| Adherence to prescribed treatment | 198 | 180 (90.9%) | 203 | 167 (82.3%) | 1.08 (1.04, 1.12) | **<0.001** | 6.8 (3.4, 10.2) |
| Adherence to diet plan | 198 | 182 (91.9%) | 203 | 167 (82.3%) | 1.07 (1.02, 1.13) | **0.007** | 6.1 (1.8, 10.4) |
| Adherence to physical activity | 198 | 181 (91.4%) | 203 | 143 (70.4%) | 1.23 (1.16, 1.30) | **<0.001** | 17.2 (12.4, 22.0) |

*\*Relative Risk was estimated via generalized linear model using Poisson regression, adjusting for age, sex, site, and time. Estimates combined all non-missing values collected at baseline and end of the study.*
*#only reported for heart failure patients.*
Adherence to prescribed treatment, diet plan and physical activity was self-reported and measured on number of days adhered in the last week and defined as $\geq 4$ days in the past week.
**Abbreviations:** LDLc = Low-density lipoprotein cholesterol; FBG = Fasting Blood Glucose; HbA1c = glycated haemoglobin; ACEi = Angiotensin-Converting Enzyme Inhibitors; ARB = Angiotensin Receptor Blocker; ARNI = Angiotensin Receptor-Neprilysin Inhibitor; CCBs = Calcium Channel Blocker, MRA=Mineralocorticoid Receptor Antagonists; SGLT2i=Sodium-Glucose Cotransporter-2 inhibitors

Figure 3 reports data on routine monitoring of risk factors, health status, and treatment satisfaction at EOS. Participants in the C-QIP arm had more frequent clinical interactions with the care providers in the past one year: mean (SD) number of outpatient clinic visits was 7.1 (4.0) vs. 2.0 (2.8) (β coefficient=2.17, 95% CI: 1.56–2.78; p<0.001), and BP checks at clinic was 7.6 (4.3) vs. 1.6 (2.9) (β=2.87, 95% CI: 2.14–3.60; p<0.001). There was small but significant improvement in the physical health T-score in the C-QIP strategy group compared to usual care group at the study end (46.9 vs 48.6; β=0.95, 95% CI: 0.04–1.85; p=0.041). Treatment satisfaction scores were also significantly higher across all domains, e.g., mean (SD) satisfaction with current treatment: 5.0 [1.3] vs. 4.5 [1.6] (β=0.51, 95% CI: 0.39–0.63; p<0.001).

**Figure 3:**
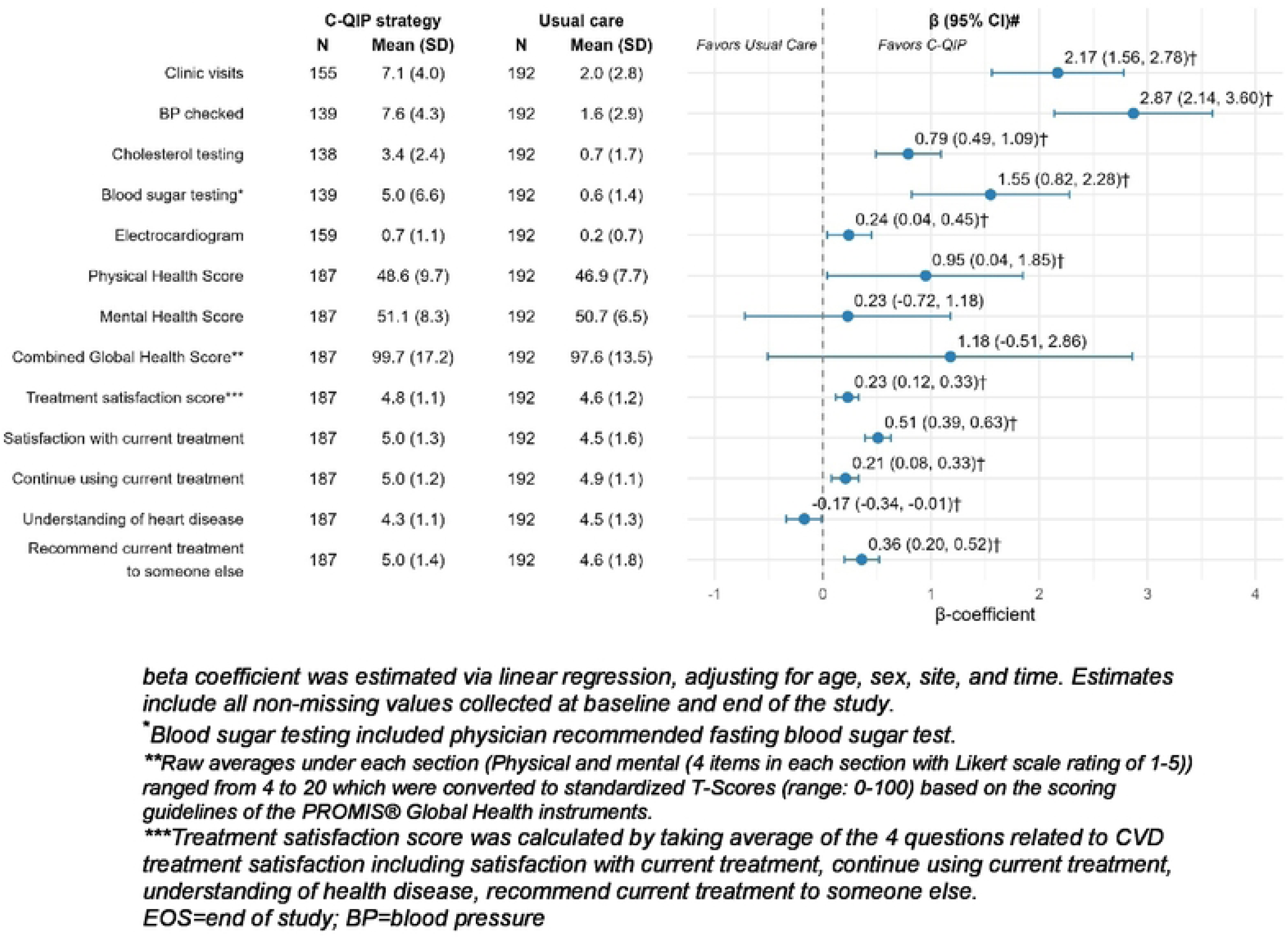
Processes of care, health status, and treatment satisfaction at EOS

Reminder systems were markedly enhanced in the C-QIP strategy arm vs usual care, phone calls after missed appointments 70.7% vs 4.4%, and calls prompting lab investigations 41.4% vs 0.5% (both p < 0.001) (**Table S.3**). Consultation time was also longer at in the C-QIP compared to usual care group (median 10 vs. 7 minutes, *p*<0.001). At trial end, family involvement in the C-QIP group was significantly higher across several self-care domains such eating healthy, exercise and quit smoking. Family support for smoking cessation was reported to be stronger in the C-QIP vs usual care arm (family joined: 7.7% vs. 0%, p=0.05; family reminded: 30.8% vs. 2.6%, p<0.001, **Table S.4:** Family support for self-care behaviours).

### Exploratory clinical measures: cardiovascular risk factors

At EOS, diastolic BP was significantly lower in the C-QIP group: 72.4 mmHg (SD 9.6) vs. 74.3 mmHg (SD 9.6); β coefficient = –1.90, 95% CI: –3.56 to –0.23; p=0.03. Systolic BP showed a small but non-significant reduction in the C-QIP vs usual care group: 118.8 mmHg (SD 15.5) vs. 121.0 mmHg (SD 15.3); β = –2.44, 95% CI: –5.07 to 0.20; p=0.07. No significant differences were observed in LDL cholesterol, HbA1c, or other risk factors (Figure 4).

**Figure 4:**
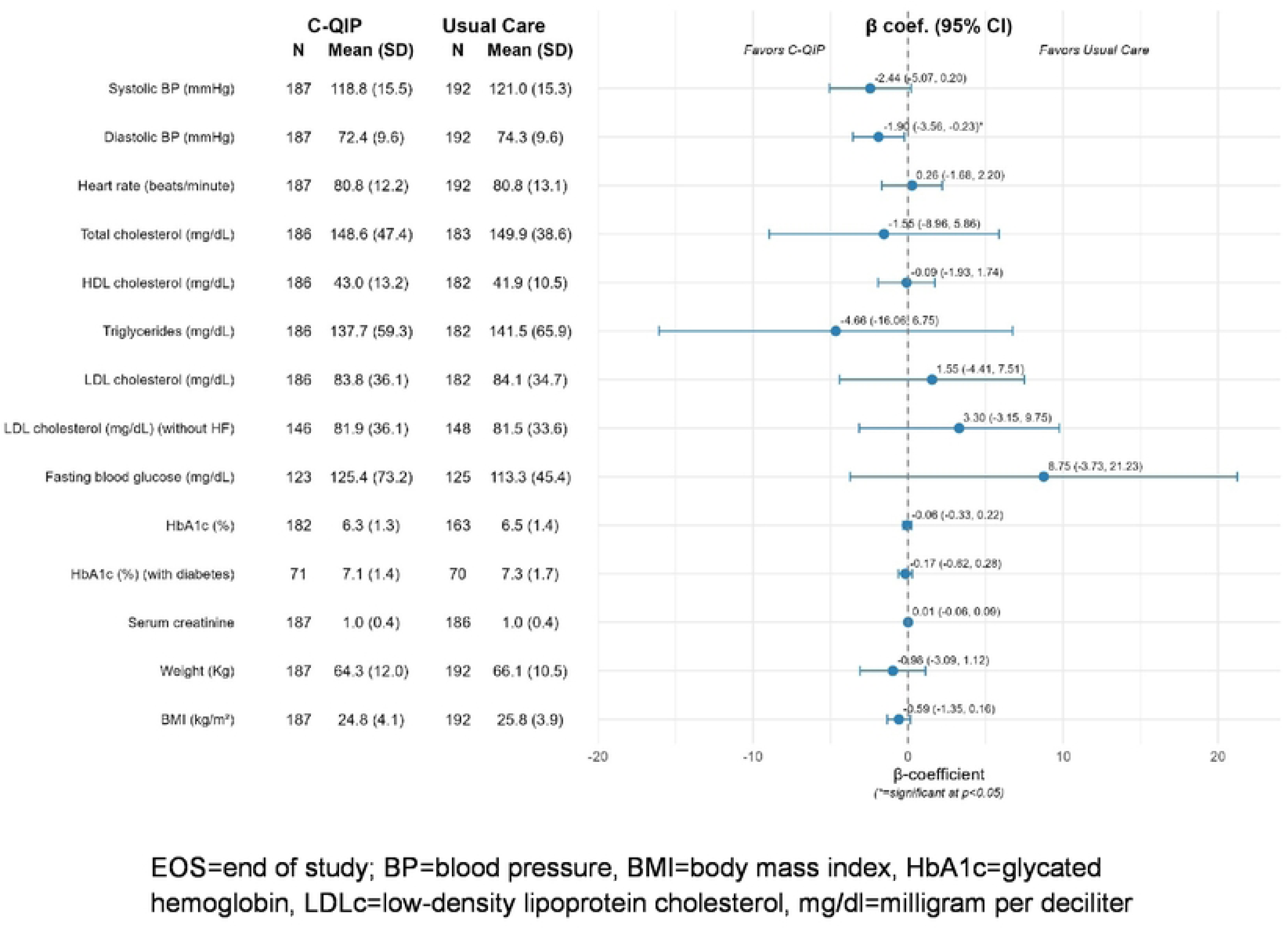
Clinical measures by treatment group at EOS

#### Sub-group analysis

No significant interactions were apparent between any of the prespecified subgroups and the treatment groups (C-QIP vs usual care arms) except private healthcare facility, and patients with stroke. (**Figure S.2:** Medication adherence by prespecified subgroups and **Figure S.3:** Systolic BP by prespecified subgroups)

#### Serious adverse events

A total of 19 deaths were reported: 9 in C-QIP and 10 in usual care arm, including 11 cardiovascular deaths and 8 non-cardiovascular deaths from diverse causes. Five hospitalizations were reported during the trial. (**Table S.5**). There were no major differences in sociodemographic or clinical characteristics between participants that completed end of study visit assessment and those lost-to-follow-up (**Table S.6**)

## Discussion

### Summary of main findings

The C-QIP trial demonstrated that a multicomponent, quality-improvement strategy for chronic CVD care was both *feasible* and *acceptable* to patients and providers. Patient recruitment, retention, and follow-up rates were high, and fidelity to the planned intervention, including delivering patient education on adherence to therapy, lifestyle advice, patient diary and VITA tool, DSS prompts review, and structured follow-up was maintained across sites. Adoption of DSS prompts by physicians varied by prompt type, yet overall uptake and user satisfaction were encouraging, highlighting the feasibility of embedding decision-support tools into routine outpatient CVD care. Patient and provider acceptability of the C-QIP strategy was high, with most respondents expressed willingness to continue using C-QIP strategy beyond trial duration. As a result, the intervention produced measurable improvements in GDMT prescription rates and self-reported adherence to medication, diet, and physical-activity. Processes of care such as timeliness of follow-up, counselling for lifestyle modification, and family engagement also improved. Exploratory analyses of CVD risk factors showed small, non-significant trends in favor of the C-QIP arm attributed to the limited power to detect between-group differences in BP, lipids, or HbA1c.

### Comparison with contemporary CVD secondary prevention trials

Compared with large-scale CVD implementation trials, C-QIP achieved similar or greater gains in implementation *processes and uptake measures* despite its resource-constrained setting. Few CVD secondary prevention trials reported implementation outcomes in a structured way. C-QIP prospectively measured feasibility, fidelity, adoption, and acceptability and demonstrated high delivery of coordinator-led counselling across visits; routine use and clinician acceptance of eDSS prompts (with variation by domain); and strong retention; and high patient/provider acceptability. Among Indian trials, ACS-QUIK (stepped-wedge cluster RCT) also paired processes of care gains with a mixed-methods process evaluation[16, 17], documenting clinician acceptability, barriers, and site-level variation, methodologically closest to C-QIP, though focused on *acute* rather than chronic care. SPREAD[18] (post-ACS, CHW-led adherence/lifestyle support) reported high visit completion and patient engagement showing evidence of *feasibility and acceptability* for task-sharing in India, but provided fewer granular fidelity or adoption metrics. CARRS Translation Trial[27] among patients with poorly controlled type 2 diabetes, demonstrated sustained program delivery in routine diabetes clinics and strong patient engagement, but again with less domain-specific reporting of adoption than C-QIP. Compared with international trials, COORDINATE-Diabetes[28] (cluster RCT across 42 US cardiology clinics implemented a clinic-level multifaceted program: care pathways, clinician education, audit/feedback, patient tools, increased concurrent use of evidence based drugs in AsCVD and type 2 diabetes patients) and BRIDGE[29] (cluster RCT of case-management, decision support, audit/feedback and patient/clinician education in primary care in Brazil) describe clinic-level education/audit-feedback performance and site engagement but provide limited, granular implementation metrics in the main study reports. NAILED-Stroke[30, 31] (nurse-led telephone follow-up with medication titration in Sweden) and STRONG-HF[32] (of protocol based high-intensity up-titration with close follow-up) implicitly demonstrate fidelity through protocol-based follow-up and titration, while CONNECT-HF[33] (cluster RCT of hospital based + post-discharge quality improvement program) underscores the challenge of scaling education/audit-feedback with heterogeneous adoption across 161 hospitals. Overall, C-QIP adds unique, LMIC-based implementation granularity (including eDSS adoption) that complements efficacy signals and bridges India’s prior acute and diabetes implementation experience into chronic CVD care.

C-QIP achieved meaningful gains in the use of GDMT: higher use of antiplatelets, statins, ACEi/ARB, β-blockers, and multi-drug GDMT. It also improved self-reported adherence to medication, diet, and physical activity. These improvements in the GDMT use and medication adherence are directionally consistent with COORDINATE-Diabetes [28] and BRIDGE trials [29]. In India, ACS-QUIK improved *acute* process-of-care measures hospital-wide[16], whereas C-QIP translates that system-thinking into the *chronic* phase with sustained gains in prescribing and adherence. SPREAD Trial[18] similarly improved post-ACS adherence and lifestyle practices with community health worker support—convergent with C-QIP’s cardiovascular care coordinator-led counselling—but without the eDSS layer that likely aided prescriber behavior in C-QIP. CARRS Translation Trial[19] showed that multicomponent strategy can improve longitudinal adherence and care processes in diabetes; C-QIP extends this experience to multiple-condition CVD (ischemic heart disease, ischemic stroke, heart failure) and demonstrates parallel improvements in medication bundles and patient behaviors. In heart failure focused implementation trials, EPIC-HF[34] (comprising of patient activation tool: 3-minute video and 1-page checklist delivered electronically before clinic consultation) and PROMPT-HF[35] (EHR based best-practice alerts) similarly moved near-term GDMT initiation or intensification. Conversely, CONNECT-HF despite scale was neutral on its composite clinical and quality endpoints, highlighting that contextual fit and implementation strength, which C-QIP emphasizes and measures, are critical for translating toolkits into clinical practice change.

C-QIP was not powered for between-group differences in BP, LDL-C, or HbA1c and showed small, non-significant trends (e.g., modest diastolic BP reduction). In contrast, trials designed around protocol-based titration to targets notably NAILED-Stroke [31] improved BP and LDL-C over 12–36 months and STRONG-HF [32] involving frequent follow-ups, GDMT up-titration with better 180-day clinical outcomes, demonstrated that dose and pace of follow-up are key to achieving CVD risk factor control. On the other hand, TEXTMEDS[36] (SMS-only) did not improve LDL-C or adherence, underscoring that single-component text-message based reminders system is insufficient. Taken together, these comparisons suggest that C-QIP’s process of care improvements are consistent with other landmark CVD trials, and that future, large hybrid trial with longer term follow-up is the logical next step to detect meaningful differences in CVD risk factors and events.

### Implications of C-QIP Trial results within the Indian context

The trial’s findings are directly relevant to India’s NP-NCD program [14], which emphasize longitudinal, team-based chronic-care management for CVD, diabetes, and hypertension. C-QIP builds on prior Indian experiences (ACS-QUIK, CARRS Translation, and SPREAD), and extends them to sustained, people-centered chronic CVD care. Demonstrating feasibility of a coordinator-supported, decision-aided model within mixed public and private settings in India, C-QIP trial supports integration of digital clinical decision support and counselling workflows into NP-NCD clinics. Importantly, C-QIP highlights the role of patient-family engagement and routine follow-up reminders, aligning with current global calls for *people-centered, integrated chronic-care models* in LMICs. Broader adoption could strengthen guideline adherence, improve continuity, and potentially reduce premature CVD mortality if implemented effectively and sustained.

### Strengths and limitations

The C-QIP trial has several notable strengths. First, it is among the few randomized implementation trials in LMICs that prospectively measured and reported core implementation outcomes: feasibility, fidelity, adoption, and acceptability alongside clinical process measures. The intervention integrated a decision-support system, structured counselling by cardiovascular care coordinators, and automated reminders, thereby demonstrating that such multicomponent strategies are feasible in routine outpatient settings across diverse public and private hospitals in India. Second, the study benefited from high recruitment, retention, and follow-up rates, reflecting the feasibility and cultural acceptability of the C-QIP strategy. Third, the multicenter implementation across varying levels of health infrastructure allowed assessment of contextual adaptability, showing that cardiovascular care coordinator–led quality improvement strategies can be integrated into existing workflows without disrupting service delivery. Fourth, the use of electronic data capture and centralized monitoring minimized data loss and enabled real-time feedback. Finally, the intervention’s grounding in prior successful models such as ACS-QUIK, SPREAD, and CARRS Translation Trial strengthens and creates a bridge from acute and CVD risk reduction to the chronic-care phase of CVD management. Collectively, these features make C-QIP a pragmatic, scalable unit for improving the quality of long-term CVD care in resource-constrained environments.

Several limitations warrant consideration. The individual-level randomization used within clinics, while pragmatic, may have led to contamination between intervention and control arms, as providers exposed to decision-support prompts could have altered care for control patients. The trial was not blinded, and several implementation measures, including provider fidelity and patient adherence, relied partly on self-report, raising the possibility of social desirability and ascertainment bias. Baseline prescription rates of evidence-based medications were already high, potentially limiting the measurable margin of improvement and attenuating effect sizes. Moreover, the trial was not powered for clinical outcomes; small differences in CVD risk factors such as diastolic BP or LDL-C must therefore be interpreted cautiously, as chance findings cannot be excluded. Finally, implementation was conducted under research-supported conditions with dedicated coordinators, and real-world rollout under programmatic conditions (e.g., under NP-NCD) may face resource and workload challenges. Despite these limitations, the trial’s consistent direction of effect across implementation measures, processes of care, and behavioral outcomes underscores the robustness of its design and provides a strong foundation for larger confirmatory hybrid effectiveness–implementation studies.

## Conclusions

The C-QIP trial demonstrated feasibility, fidelity, and acceptability of a multicomponent CVD care strategy across Indian outpatient settings. The intervention achieved significant improvements in guideline-directed prescribing, patient adherence, and care processes, with strong participant retention and no excess in adverse events. While CVD risk factor changes were modest, the process and implementation gains provide the empirical foundation for future large-scale, outcomes-driven hybrid effectiveness–implementation trial to reduce CVD morbidity and mortality.

## Data Availability

The data underlying the results presented in the study are available from the corresponding author: Kavita Singh

## Acknowledgements

We acknowledge the patients and teams at each of the four participating hospitals in C-QIP Trial. We acknowledge the work of the software company, QuadOne Clinion, for developing the study database. We acknowledge the support of the Data Safety and Monitoring Board committee for supervision.

**DSMB members:** Dr. Nitish Naik (Chair), Dr. PP Mohanan, Dr. Salim Virani, Dr. Ratna Devi, Dr. Laurent Billot.

**Participating investigator sites**: (1) All India Institute of Medical Sciences (AIIMS), New Delhi, India: PI: Dr. Ambuj Roy, SC & RA: Kamar Ali, CCC: Mohit Sharma; (2) Sir Ganga Ram Hospital, New Delhi, India: PI: Dr. JPS Sawhney, Co-I: Dr. Kushal Madan, Dr. Kavita Tyagi, SC: Ajeet Nanda, CCC: Bhumika Jalutharia, RA: Tanvi Dhiman; (3) GB Pant Hospital, New Delhi, India: PI: Dr. Girish MP, Co-I: Dr. Mohit Gupta, SC & RA: Aarti Gupta, CCC: Manasvi Shukla; (4) SDM College of Medical Sciences and Hospital, Karnataka, India: PI: Dr. Kiran Aithal, Co-I: Dr. Satish G. Patil, Dr Sheela S. Kudari, Dr Bharati A. Gadad, Dr Vitthal Khode CCC: Laxmi Ramchandra Patil, RA: Pooja Katti.

## Data availability statement

KS and DK have access to study data set and statistical code. Any request for data sharing should be addressed to the corresponding author (KS).

---

**Figure S.1.**
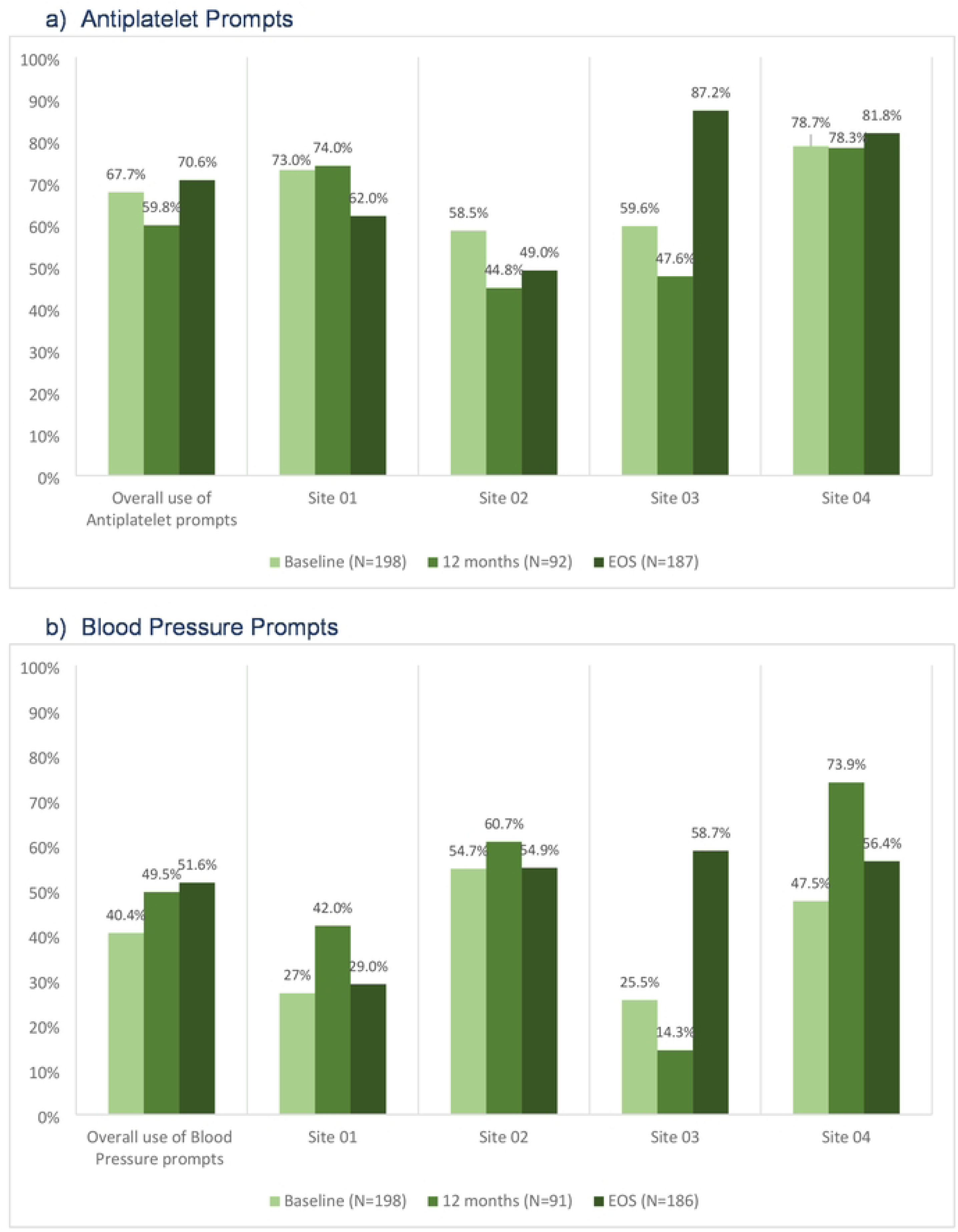
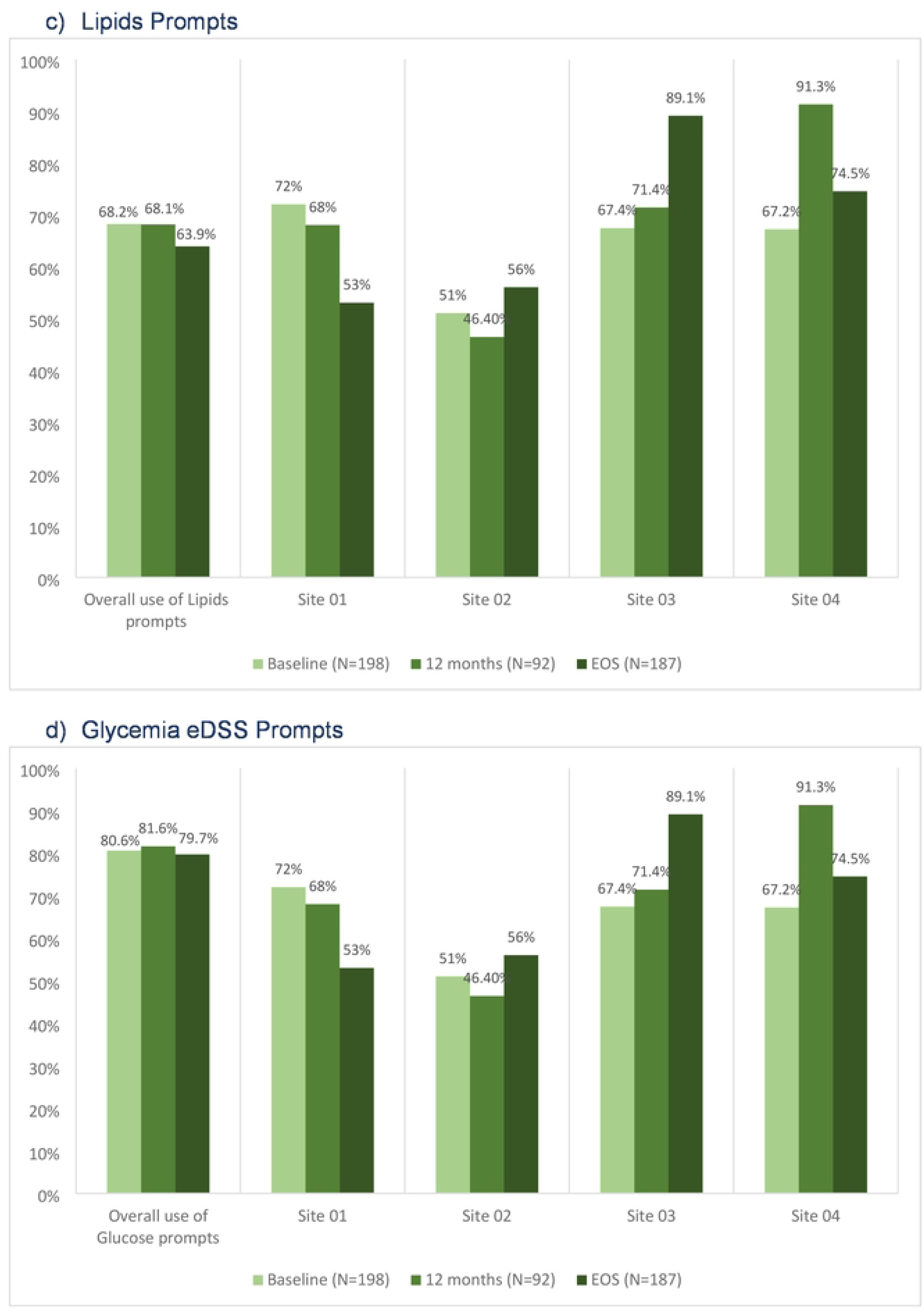
Adoption of electronic decision support software (eDSS) prompts review and acceptance by physicians: overall and by site.

**Figure S.2:**
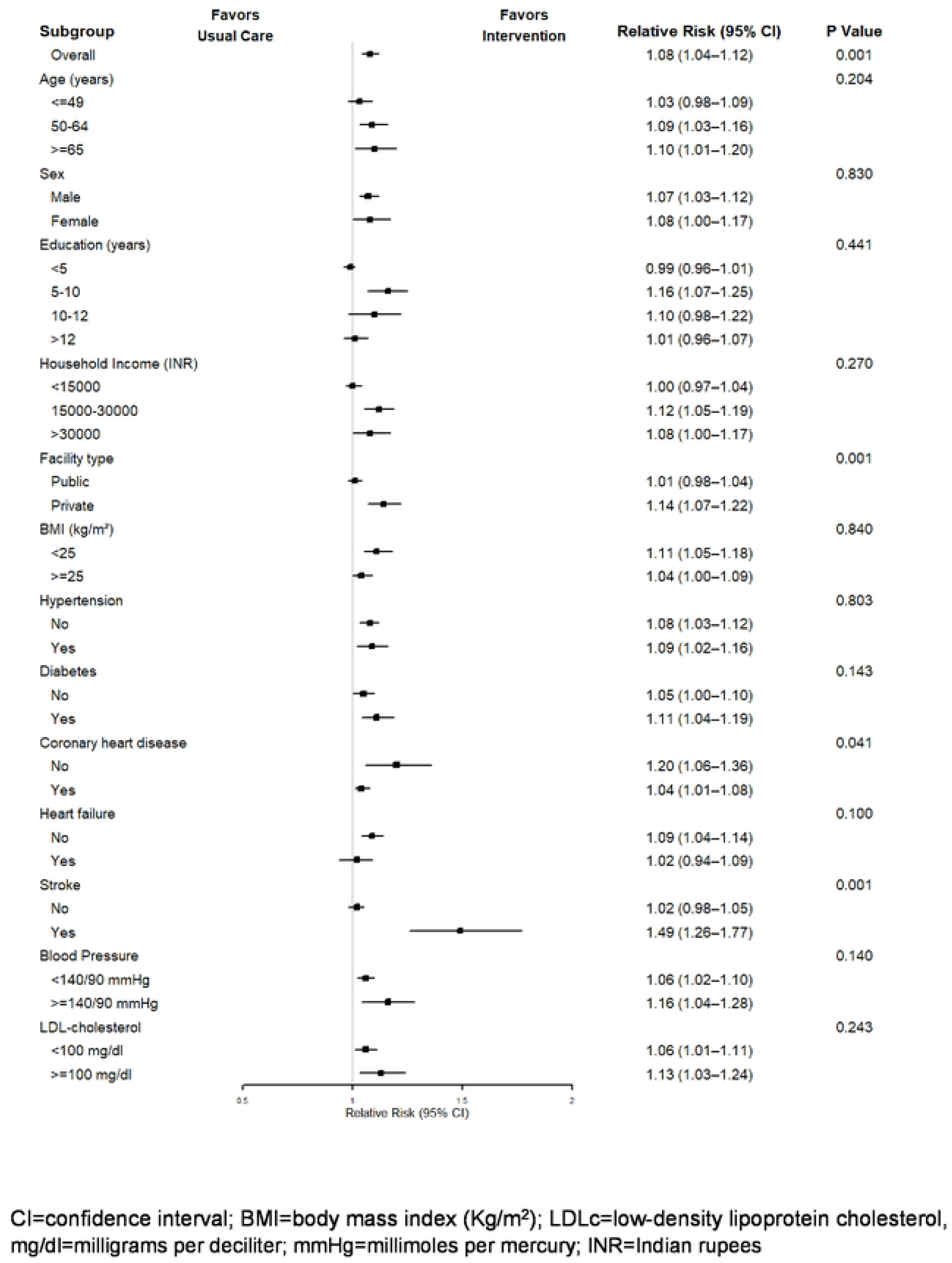
Self-reported medication adherence at EOS by prespecified sub-groups

**Figure S.3:**
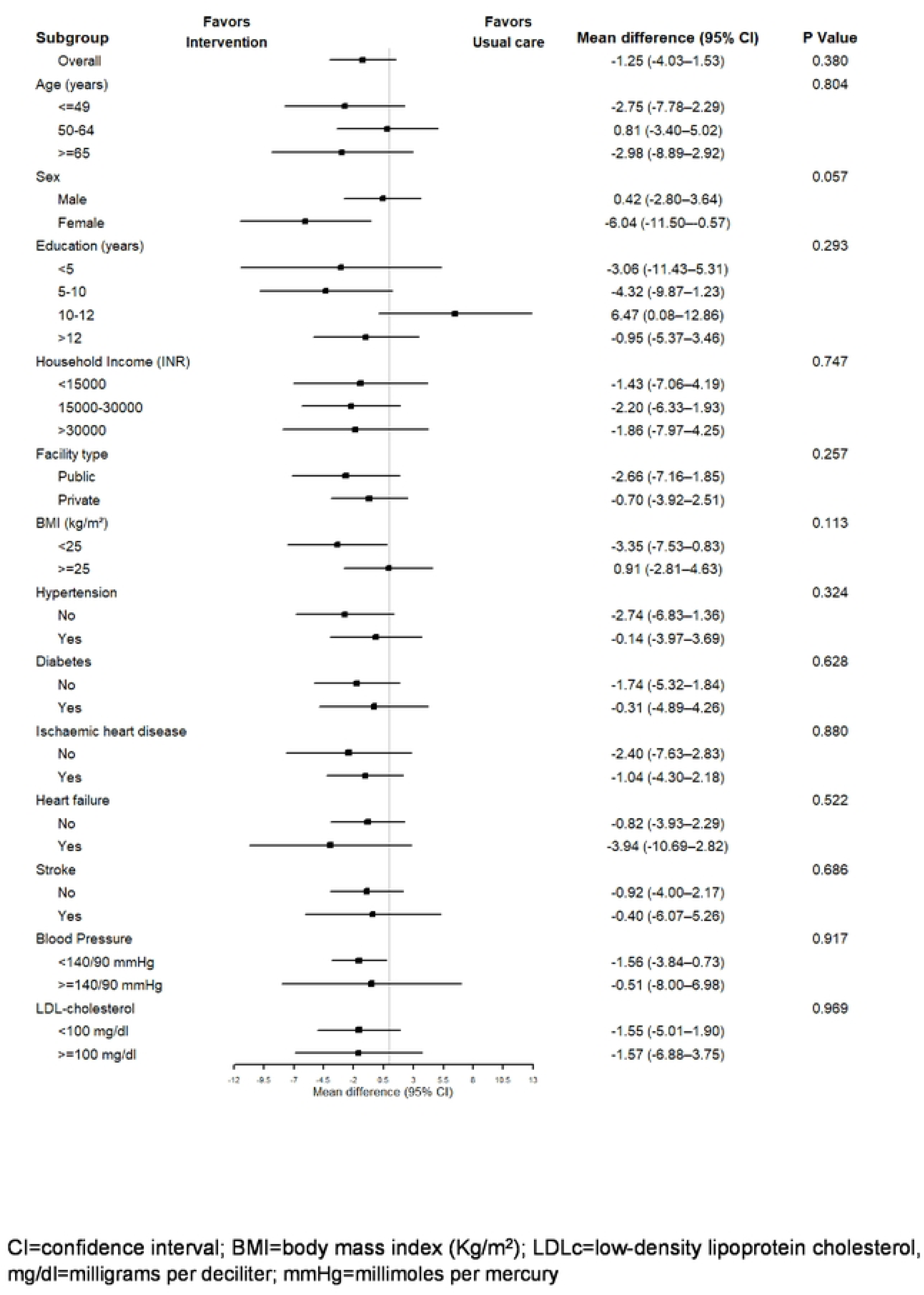
Mean systolic blood pressure reduction at end-of-study by prespecified subgroups

**Table S.1.** : Fidelity of C-QIP Strategy: Self-reported cardiovascular care coordinator delivered counselling of lifestyle modifications for intervention arm participants.

|  | Baseline<br>(N=198) | 12 months<br>follow-up<br>(N=114) | EOS<br>(N=197) |
| --- | --- | --- | --- |
| **Participant counselled for lifestyle modifications** | **197 (99.5%)** | **91 (79.8%)** | **187 (94.4%)** |
| Healthy diet | 197 (100%) | 91 (100%) | 186 (99.5%) |
| Exercise | 197 (100%) | 91 (100%) | 184 (98.4%) |
| Smoking cessation<br>(If said yes/quit to smoking) | 24 (68.6%)<br>(n=35) | 6 (100%)<br>(n=6) | 9 (60%)<br>(n=15) |
| Avoidance of stress | 159 (80.7%) | 82 (90.1%) | 170 (90.9%) |
| Abstinence/ limited use of alcohol<br>(If said yes to alcohol) | 10 (83.3%)<br>(n=12) | 10 (83.3%)<br>(n=12) | 2 (33.3%)<br>(n=6) |
| Home monitoring of blood pressure | 159 (80.7%) | 83 (91.2%) | 169 (90.4%) |

**Table S.2.** : Feasibility of eDSS recommended intermediate follow-up visits completion for intervention arm participants.

| Intermediate Visit (IV) | Expected | Completed |
| --- | --- | --- |
| **Between baseline and annual visits** |  |  |
| IV 2.1 | 195 | 193 |
| IV 2.2 | 193 | 188 |
| IV 2.3 | 188 | 180 |
| IV 2.4 | 151 | 117 |
| IV 2.5 | 97 | 62 |
| IV 2.6 | 53 | 31 |
| IV 2.7 | 26 | 13 |
| IV 2.8 | 13 | 3 |
| IV 2.9 | 3 | 1 |
| **Post annual visits** |  |  |
| IV 3.1 | 66 | 55 |
| IV 3.2 | 41 | 33 |
| IV 3.3 | 17 | 7 |
| IV 3.4 | 4 | 1 |
| IV 3.5 | 1 | 1 |

IV=Intermediate visits; eDSS=electronic decision support system
*The table above shows the expected and actual number of completed intermediate visits between the baseline and annual visits, and visits completed between the first annual visit and end of the study visit.*
*Average follow-up visit duration between intermediate visits was approximately 3 months or as recommended per the eDSS prompts for chronic care of patients with ischemic heart disease, ischemic stroke or heart failure regardless of ejection fraction.*

**Table S.3.** : Self-reported timeliness of care and type of reminders received.

|  | Baseline |  | End of Study (EOS) |  | p-value* |
| --- | --- | --- | --- | --- | --- |
|  | C-QIP strategy<br>(N=198) | Usual care<br>(N=203) | C-QIP strategy<br>(N=187) | Usual care<br>(N=192) |  |
| Total time spend for lab tests, include waiting (in minutes; Median, IQR) | 30 (15, 45) | 20 (15, 35) | 20 (15, 40) | 30 (15, 60) | <0.001 |
| Total time spend waiting for consultation (in minutes; Median, IQR) | 20 (10, 40) | 20 (10, 60) | 15 (10, 25) | 20 (10, 40) | 0.13 |
| Total time spend in-person with doctor (in minutes; Median, IQR) | 9 (5, 10) | 8 (5, 10) | 10 (5, 10) | 7 (5, 10) | <0.001 |
| **Type of reminders received** |  |  |  |  |  |
| Verbal reminder at current visit | 147 (74.2%) | 143 (70.4%) | 130 (65.7%) | 134 (66%) | 0.95 |
| Phone call for upcoming appointment | 56 (28.3%) | 71 (35%) | 93 (47%) | 57 (28.1%) | <0.001 |
| Phone call if missed appointment | 43 (21.7%) | 34 (16.8%) | 140 (70.7%) | 9 (4.4%) | <0.001 |
| Phone call to go for lab investigations | 13 (6.6%) | 9 (4.4%) | 82 (41.4%) | 1 (0.5%) | <0.001 |

\*p-value is for the difference between the two groups at end of study

**Table S.4.** : Self-reported family support at baseline and end of study.

| Family has helped the participant to | Baseline |  | End of Study (EOS) |  | p-value* |
| --- | --- | --- | --- | --- | --- |
|  | C-QIP strategy (N=198) | Usual care (N=203) | C-QIP strategy (N=198) | Usual care (N=203) |  |
| Eat healthy | 182 (91.9%) | 146 (71.9%) | 187 (94.4%) | 138 (68%) |  |
| Exercise | 150 (75.8%) | 126 (62.1%) | 185 (93.4%) | 132 (65%) | 0.06 |
| Quit smoking<br>(If said yes/quit to smoking) | N=35<br>14 (40%) | N=26<br>7 (15.6%) | N=45<br>9 (34.6%) | N=38<br>1 (2.6%) | <0.001 |
| Family reminds participant to |  |  |  |  |  |
| Eat healthy | 176 (88.9%) | 141 (69.5%) | 187 (94.4%) | 138 (68%) |  |
| Exercise | 157 (79.3%) | 129 (63.6%) | 181 (91.4%) | 127 (62.6%) | 0.06 |
| Quit smoking<br>(If said yes/quit to smoking) | N=35<br>14 (40%) | N=26<br>8 (17.8%) | N=45<br>8 (30.8%) | N=38<br>1 (2.6%) | <0.001 |
| Family joins with the participant to |  |  |  |  |  |
| Eat healthy | 170 (85.9%) | 136 (67%) | 186 (93.9%) | 137 (67.5%) | 0.39 |
| Exercise | 103 (52%) | 75 (37%) | 127 (64.1%) | 96 (47.3%) | 0.68 |
| Quit smoking<br>(If said yes/quit to smoking) | N=35<br>7 (20%) | N=26<br>2 (4.4%) | N=45<br>2 (7.7%) | N=38<br>- | 0.05 |

\*p-value is for the difference between the two groups at EOS

**Table S.5.** : Serious adverse events.

| Event description | Total events reported in the trial |  |  |
| --- | --- | --- | --- |
|  | Total | C-QIP strategy | Usual care |
| **Deaths (all cause)** | **19** | **9** | **10** |
| Cardiovascular deaths | 11 | 6 | 5 |
| Other deaths* | 8 | 3 | 5 |
| Death followed by severe illness |  |  | 3 |
| Road traffic accident |  |  | 1 |
| Cancer complications |  | 1 |  |
| Deteriorating health |  | 1 |  |
| Post-operation complications |  | 1 |  |
| Difficulty in breathing |  |  | 1 |
| **Hospitalizations** | **5** | **5** | **0** |
| Angina on exertion | 2 | 2 |  |
| Chest pain | 1 | 1 |  |
| Anteroseptal Myocardial Infarction | 1 | 1 |  |
| Planned angiography | 1 | 1 |  |

**Table S.6.** : Comparison of baseline characteristics of participants who completed the EOS visit versus participants who did not complete EOS visit.

|  | Completed EOS visit | Lost to follow up | p-value |
| --- | --- | --- | --- |
|  | N=379 | N=22 |  |
| Age (years; mean, SD) | 57.1 (11.7) | 62.7 (11.0) | 0.03 |
| Male, n (%) | 277 (73.1%) | 16 (72.7%) | 0.97 |
| Education, (years; mean, SD) | 10.7 (5.6) | 11.6 (4.3) | 0.44 |
| Household Income (monthly)<br>Indian rupees, median (IQR) | 20000 (10000, 30000) | 27500 (20000, 30000) | 0.05 |
| Systolic BP (mm Hg; mean, SD) | 127.4 (18.1) | 134.5 (19.9) | 0.07 |
| Diastolic BP (mm Hg; mean, SD) | 76.2 (11.4) | 78.8 (14.3) | 0.30 |
| Total cholesterol (mg/dL; mean, SD) | 144.3 (44.2)<br>(n=370) | 149.0 (37.7) (n=22) | 0.63 |
| LDLc (mg/dL; mean, SD) | 80.2 (37.4)<br>(n=367) | 84.7 (35.2) (n=21) | 0.59 |
| FBG (mg/dL; mean, SD) | 123.0 (55.2)<br>(n=182) | 87.6 (23.6)<br>(n=4) | 0.20 |
| HbA1c (%; mean, SD) | 6.7 (1.6)<br>(n=225) | 8.0 (1.7)<br>(n=7) | 0.03 |
| **Medical history** |  |  |  |
| Coronary heart disease, n (%) | 286 (75.5%) | 10 (45.5%) | 0.002 |
| Heart failure, n (%) | 76 (20.1%) | 6 (27.3%) | 0.41 |
| Ischemic stroke, n (%) | 65 (17.2%) | 9 (40.9%) | 0.005 |
| Hypertension, n (%) | 178 (47.0%) | 15 (68.2%) | 0.05 |
| Diabetes, n (%) | 155 (40.9%) | 13 (59.1%) | 0.09 |
| **Medications use** |  |  |  |
| Antiplatelet | 336 (88.7%) | 15 (68.2%) | 0.005 |
| Lipid lowering (Statin) | 330 (87.1%) | 14 (63.6%) | 0.002 |
| ACEi/ARB | 227 (59.9%) | 12 (54.5%) | 0.62 |
| ARNI (n=82; heart failure patients) | 24 (32%) | 1 (17%) | 0.44 |
| Beta-blockers | 269 (71.0%) | 15 (68.2%) | 0.78 |
| Calcium Channel Blockers | 64 (16.9%) | 7 (31.8%) | 0.07 |
| Diuretics | 29 (7.7%) | 1 (4.5%) | 0.59 |
| Did participant forget to take medications (in last 1 month)? | 56 (14.8%) | 3 (13.6%) | 0.96 |
| On feeling worse does the participant stops medication on his/her own? | 19 (5.0%) | 0 (0.0%) | 0.28 |

\*EOS=End of study visit; SD=standard deviation, mg/dl=milligrams per deciliter, BP=blood pressure

## References

1. Chong B, Jayabaskaran J, Jauhari SM, Chan SP, Goh R, Kueh MTW, et al. Global burden of cardiovascular diseases: projections from 2025 to 2050. Eur J Prev Cardiol. 2024. Epub 20240913. doi: 10.1093/eurjpc/zwae281. PubMed PMID: 39270739.

2. Collaborators GBDCD. Global, Regional, and National Burden of Cardiovascular Diseases and Risk Factors in 204 Countries and Territories, 1990-2023. J Am Coll Cardiol. 2025. Epub 20250924. doi: 10.1016/j.jacc.2025.08.015. PubMed PMID: 40990886.

3. Leong DP, Yusuf R, Iqbal R, Avezum A, Yusufali A, Rosengren A, et al. The burden of cardiovascular events according to cardiovascular risk profile in adults from high-income, middle-income, and low-income countries (PURE): a cohort study. Lancet Glob Health. 2025;13(8):e1406–e14. doi: 10.1016/S2214-109X(25)00155-X. PubMed PMID: 40712612.

4. Walker IF, Garbe F, Wright J, Newell I, Athiraman N, Khan N, Elsey H. The Economic Costs of Cardiovascular Disease, Diabetes Mellitus, and Associated Complications in South Asia: A Systematic Review. Value Health Reg Issues. 2018;15:12–26. Epub 20170703. doi: 10.1016/j.vhri.2017.05.003. PubMed PMID: 29474174.

5. Wood DA. WHF Roadmap on Secondary Prevention of CVD. Glob Heart. 2024;19(1):9. Epub 20240122. doi: 10.5334/gh.1294. PubMed PMID: 38273997; PubMed Central PMCID: PMCPMC10809851.

6. Tabak RG, Kandula NR, Angell SY, Brewer LC, Grandner M, Hayman LL, et al. Implementation of Evidence-Based Behavioral Interventions for Cardiovascular Disease Prevention in Community Settings: A Scientific Statement From the American Heart Association. Circulation. 2025. Epub 20250804. doi: 10.1161/CIR.0000000000001349. PubMed PMID: 40755303.

7. Yoo SGK, Chung GS, Bahendeka SK, Sibai AM, Damasceno A, Farzadfar F, et al. Aspirin for Secondary Prevention of Cardiovascular Disease in 51 Low-, Middle-, and High-Income Countries. JAMA. 2023;330(8):715–24. doi: 10.1001/jama.2023.12905. PubMed PMID: 37606674; PubMed Central PMCID: PMCPMC10445202.

8. Yusuf S, Islam S, Chow CK, Rangarajan S, Dagenais G, Diaz R, et al. Use of secondary prevention drugs for cardiovascular disease in the community in high-income, middle-income, and low-income countries (the PURE Study): a prospective epidemiological survey. Lancet. 2011;378(9798):1231–43. Epub 20110826. doi: 10.1016/S0140-6736(11)61215-4. PubMed PMID: 21872920.

9. Joseph P, Avezum A, Ramasundarahettige C, Mony PK, Yusuf R, Kazmi K, et al. Secondary Prevention Medications in 17 Countries Grouped by Income Level (PURE): A Prospective Cohort Study. J Am Coll Cardiol. 2025;85(5):436–47. doi: 10.1016/j.jacc.2024.10.121. PubMed PMID: 39909677.

10. Mulure N, Hewadmal H, Khan Z. Assessing Barriers to Primary Prevention of Cardiovascular Diseases in Low and Middle-Income Countries: A Systematic Review. Cureus. 2024;16(7):e65516. Epub 20240727. doi: 10.7759/cureus.65516. PubMed PMID: 39188440; PubMed Central PMCID: PMCPMC11346380.

11. Agyemang C, van den Born BJ. Limited access to CVD medicines in low-income and middle-income countries: poverty is at the heart of the matter. Lancet Glob Health. 2018;6(3):e234–e5. doi: 10.1016/S2214-109X(18)30048-2. PubMed PMID: 29433655.

12. Mendis S, Graham I. Prevention and control of cardiovascular disease in “real-world” settings: sustainable implementation of effective policies. Front Cardiovasc Med. 2024;11:1380809. Epub 20241119. doi: 10.3389/fcvm.2024.1380809. PubMed PMID: 39628553; PubMed Central PMCID: PMCPMC11611850.

13. Schwalm JD, McKee M, Huffman MD, Yusuf S. Resource Effective Strategies to Prevent and Treat Cardiovascular Disease. Circulation. 2016;133(8):742–55. doi: 10.1161/CIRCULATIONAHA.115.008721. PubMed PMID: 26903017; PubMed Central PMCID: PMCPMC4766731.

14. National Programme for prevention and control of NCDs (NP-NCD). Ministry of Health and Family Welfare, Government of India 2023 [cited 2025 10 November]. Available from: https://ncd.nhp.gov.in/ncdlandingassets/aboutus.html.

15. Meghana RVNSP, Singh J, Huda RK. Challenges, barriers, and facilitators in implementing digital health initiatives for NP-NCD management in India’s public health system: A scoping review. Clinical Epidemiology and Global Health. 2025;35. doi: 10.1016/j.cegh.2025.102137.

16. Huffman MD, Mohanan PP, Devarajan R, Baldridge AS, Kondal D, Zhao L, et al. Effect of a Quality Improvement Intervention on Clinical Outcomes in Patients in India With Acute Myocardial Infarction: The ACS QUIK Randomized Clinical Trial. JAMA. 2018;319(6):567–78. doi: 10.1001/jama.2017.21906. PubMed PMID: 29450524; PubMed Central PMCID: PMCPMC5838631.

17. Singh K, Devarajan R, Mohanan PP, Baldridge AS, Kondal D, Victorson DE, et al. Implementation and acceptability of a heart attack quality improvement intervention in India: a mixed methods analysis of the ACS QUIK trial. Implement Sci. 2019;14(1):12. Epub 20190206. doi: 10.1186/s13012-019-0857-7. PubMed PMID: 30728053; PubMed Central PMCID: PMCPMC6364470.

18. Xavier D, Gupta R, Kamath D, Sigamani A, Devereaux PJ, George N, et al. Community health worker-based intervention for adherence to drugs and lifestyle change after acute coronary syndrome: a multicentre, open, randomised controlled trial. Lancet Diabetes Endocrinol. 2016;4(3):244–53. Epub 20160206. doi: 10.1016/S2213-8587(15)00480-5. PubMed PMID: 26857999.

19. Ali MK, Singh K, Kondal D, Devarajan R, Patel SA, Menon VU, et al. Effect of a multicomponent quality improvement strategy on sustained achievement of diabetes care goals and macrovascular and microvascular complications in South Asia at 6.5 years follow-up: Post hoc analyses of the CARRS randomized clinical trial. PLoS Med. 2024;21(6):e1004335. Epub 20240603. doi: 10.1371/journal.pmed.1004335. PubMed PMID: 38829880; PubMed Central PMCID: PMCPMC11198027.

20. Singh K, Johnson L, Devarajan R, Shivashankar R, Sharma P, Kondal D, et al. Acceptability of a decision-support electronic health record system and its impact on diabetes care goals in South Asia: a mixed-methods evaluation of the CARRS trial. Diabet Med. 2018;35(12):1644–54. Epub 20180919. doi: 10.1111/dme.13804. PubMed PMID: 30142228.

21. Singh K, Nikhare K, Gandral M, Aithal K, Patil SG, Mp G, et al. Rationale, Design and Baseline Characteristics of a Randomized Controlled Trial of a Cardiovascular Quality Improvement Strategy in India: The C-QIP Trial. Am Heart J. 2024;276:83–98. Epub 20240720. doi: 10.1016/j.ahj.2024.07.008. PubMed PMID: 39033994; PubMed Central PMCID: PMCPMC12423933.

22. Singh K, Bawa VS, Venkateshmurthy NS, Gandral M, Sharma S, Lodhi S, et al. Assessment of Studies of Quality Improvement Strategies to Enhance Outcomes in Patients With Cardiovascular Disease. JAMA Netw Open. 2021;4(6):e2113375. Epub 20210601. doi: 10.1001/jamanetworkopen.2021.13375. PubMed PMID: 34125220; PubMed Central PMCID: PMCPMC8204210.

23. Singh K, Huffman MD, Johnson LCM, Tandon N, Prabhakaran D, Mendenhall E. Collaborative Quality Improvement Strategy in Secondary Prevention of Cardiovascular Disease in India: Findings from a Multi-Stakeholder, Qualitative Study using Consolidated Framework for Implementation Research (CFIR). Glob Heart. 2022;17(1):72. Epub 20221011. doi: 10.5334/gh.1161. PubMed PMID: 36382156; PubMed Central PMCID: PMCPMC9562780.

24. Singh K, Joshi A, Venkateshmurthy NS, Rahul R, Huffman MD, Tandon N, Prabhakaran D. A Delphi Study to Prioritize Evidence-Based Strategies for Cardiovascular Disease Care in India. Glob Implement Res Appl. 2023:1–12. Epub 20230605. doi: 10.1007/s43477-023-00087-2. PubMed PMID: 37363377; PubMed Central PMCID: PMCPMC10240122.

25. Billingham SA, Whitehead AL, Julious SA. An audit of sample sizes for pilot and feasibility trials being undertaken in the United Kingdom registered in the United Kingdom Clinical Research Network database. BMC Med Res Methodol. 2013;13:104. Epub 20130820. doi: 10.1186/1471-2288-13-104. PubMed PMID: 23961782; PubMed Central PMCID: PMCPMC3765378.

26. Totton N, Lin J, Julious S, Chowdhury M, Brand A. A review of sample sizes for UK pilot and feasibility studies on the ISRCTN registry from 2013 to 2020. Pilot Feasibility Stud. 2023;9(1):188. Epub 20231121. doi: 10.1186/s40814-023-01416-w. PubMed PMID: 37990337; PubMed Central PMCID: PMCPMC10662929.

27. Johnson LCM, Nikhare K, Jaganathan S, Ali MK, Narayan KMV, Prabhakaran D, et al. Stakeholder Perspectives regarding the Acceptability and Sustainability of a Multi-component Diabetes Care Strategy in South Asia: a longitudinal qualitative analysis. Glob Implement Res Appl. 2022;2(4):350–60. Epub 20221019. doi: 10.1007/s43477-022-00060-5. PubMed PMID: 37745272; PubMed Central PMCID: PMCPMC10516368.

28. Pagidipati NJ, Nelson AJ, Kaltenbach LA, Leyva M, McGuire DK, Pop-Busui R, et al. Coordinated Care to Optimize Cardiovascular Preventive Therapies in Type 2 Diabetes: A Randomized Clinical Trial. JAMA. 2023;329(15):1261–70. doi: 10.1001/jama.2023.2854. PubMed PMID: 36877177; PubMed Central PMCID: PMCPMC9989955.

29. Machline-Carrion MJ, Soares RM, Damiani LP, Campos VB, Sampaio B, Fonseca FH, et al. Effect of a Multifaceted Quality Improvement Intervention on the Prescription of Evidence-Based Treatment in Patients at High Cardiovascular Risk in Brazil: The BRIDGE Cardiovascular Prevention Cluster Randomized Clinical Trial. JAMA Cardiol. 2019;4(5):408–17. doi: 10.1001/jamacardio.2019.0649. PubMed PMID: 30942842; PubMed Central PMCID: PMCPMC6537802.

30. Irewall AL, Ogren J, Bergstrom L, Laurell K, Soderstrom L, Mooe T. Nurse-Led, Telephone-Based, Secondary Preventive Follow-Up after Stroke or Transient Ischemic Attack Improves Blood Pressure and LDL Cholesterol: Results from the First 12 Months of the Randomized, Controlled NAILED Stroke Risk Factor Trial. PLoS One. 2015;10(10):e0139997. Epub 20151016. doi: 10.1371/journal.pone.0139997. PubMed PMID: 26474055; PubMed Central PMCID: PMCPMC4608694.

31. Ogren J, Irewall AL, Soderstrom L, Mooe T. Long-term, telephone-based follow-up after stroke and TIA improves risk factors: 36-month results from the randomized controlled NAILED stroke risk factor trial. BMC Neurol. 2018;18(1):153. Epub 20180921. doi: 10.1186/s12883-018-1158-5. PubMed PMID: 30241499; PubMed Central PMCID: PMCPMC6148791.

32. Mebazaa A, Davison B, Chioncel O, Cohen-Solal A, Diaz R, Filippatos G, et al. Safety, tolerability and efficacy of up-titration of guideline-directed medical therapies for acute heart failure (STRONG-HF): a multinational, open-label, randomised, trial. Lancet. 2022;400(10367):1938–52. Epub 20221107. doi: 10.1016/S0140-6736(22)02076-1. PubMed PMID: 36356631.

33. DeVore AD, Granger BB, Fonarow GC, Al-Khalidi HR, Albert NM, Lewis EF, et al. Effect of a Hospital and Postdischarge Quality Improvement Intervention on Clinical Outcomes and Quality of Care for Patients With Heart Failure With Reduced Ejection Fraction: The CONNECT-HF Randomized Clinical Trial. JAMA. 2021;326(4):314–23. doi: 10.1001/jama.2021.8844. PubMed PMID: 34313687; PubMed Central PMCID: PMCPMC8317015.

34. Allen LA, Venechuk G, McIlvennan CK, Page RL, 2nd, Knoepke CE, Helmkamp LJ, et al. An Electronically Delivered Patient-Activation Tool for Intensification of Medications for Chronic Heart Failure With Reduced Ejection Fraction: The EPIC-HF Trial. Circulation. 2021;143(5):427–37. Epub 20201117. doi: 10.1161/CIRCULATIONAHA.120.051863. PubMed PMID: 33201741; PubMed Central PMCID: PMCPMC7855616.

35. Ghazi L, Yamamoto Y, Riello RJ, Coronel-Moreno C, Martin M, O’Connor KD, et al. Electronic Alerts to Improve Heart Failure Therapy in Outpatient Practice: A Cluster Randomized Trial. J Am Coll Cardiol. 2022;79(22):2203–13. Epub 20220403. doi: 10.1016/j.jacc.2022.03.338. PubMed PMID: 35385798.

36. Chow CK, Klimis H, Thiagalingam A, Redfern J, Hillis GS, Brieger D, et al. Text Messages to Improve Medication Adherence and Secondary Prevention After Acute Coronary Syndrome: The TEXTMEDS Randomized Clinical Trial. Circulation. 2022;145(19):1443–55. Epub 20220509. doi: 10.1161/CIRCULATIONAHA.121.056161. PubMed PMID: 35533220.

